# COVID-19 Chest X-Ray Image Classification Using Deep Learning

**DOI:** 10.1101/2021.07.15.21260605

**Authors:** Gunther Correia Bacellar, Mallikarjuna Chandrappa, Rajlakshman Kulkarni, Soumava Dey

## Abstract

The rise of the coronavirus disease 2019 (COVID-19) pandemic has made it necessary to improve existing medical screening and clinical management of this disease. While COVID-19 patients are known to exhibit a variety of symptoms, the major symptoms include fever, cough, and fatigue. Since these symptoms also appear in pneumonia patients, this creates complications in COVID-19 detection especially during the flu season. Early studies identified abnormalities in chest X-ray images of COVID-19 infected patients that could be beneficial for disease diagnosis. Therefore, chest X-ray image-based disease classification has emerged as an alternative to aid medical diagnosis. However, manual detection of COVID-19 from a set of chest X-ray images comprising both COVID-19 and pneumonia cases is cumbersome and prone to human error. Thus, artificial intelligence techniques powered by deep learning algorithms, which learn from radiography images and predict presence of COVID-19 have potential to enhance current diagnosis process. Towards this purpose, here we implemented a set of deep learning pre-trained models such as ResNet, VGG, Inception and EfficientNet in conjunction with developing a computer vision AI system based on our own convolutional neural network (CNN) model: Deep Learning in Healthcare (DLH)-COVID. All these CNN models cater to image classification exercise. We used publicly available resources of 6,432 images and further strengthened our model by tuning hyperparameters to provide better generalization during the model validation phase. Our final DLH-COVID model yielded the highest accuracy of 96% in detection of COVID-19 from chest X-ray images when compared to images of both pneumonia-affected and healthy individuals. Given the practicality of acquiring chest X-ray images by patients, we also developed a web application (link: https://toad.li/xray) based on our model to directly enable users to upload chest X-ray images and detect the presence of COVID-19 within a few seconds. Taken together, here we introduce a state-of-the-art artificial intelligence-based system for efficient COVID-19 detection and a user-friendly application that has the capacity to become a rapid COVID-19 diagnosis method in the near future.

## 1. INTRODUCTION

The COVID-19 is a viral infection that causes severe respiratory illness ranging from common cold to life threating diseases like Severe Acute Respiratory Syndrome (SARS) and Middle East Respiratory Syndrome (MERS). According to reports from the World Health Organization (WHO), major symptoms of COVID-19 are similar to that of common flu: fever, tiredness, dry cough, shortness of breath, aches and sore throat [1,2]. The similarities between COVID-19 and flu symptoms causes difficulties in detection of the coronavirus at early stages. It was found in some patients that the coronavirus, like other viruses and bacteria, also causes pneumonia, and the treatment for coronavirus induced pneumonia is different from other types of pneumonia. Moreover, bacterial pneumonia infected patients require antibiotic treatment whereas viral pneumonia cases can be treated by intensive care [3]. Therefore, accurate and timely diagnosis of COVID-19 induced pneumonia is very important to save human lives as well as curbing the pandemic outbreak across the world.

The WHO approved method of testing COVID-19 is the reverse transcription polymerase chain reaction (RT-PCR) where short sequences of RNA are analyzed to detect presence of coronavirus [4]. However, following challenges are potential barriers to facilitate COVID-19 detection using current methodologies: (1) Negative results from RT-PCR do not rule out the possibility of a person infected with COVID-19. This requires the need for further assessment to confirm false negative cases [5,6]. (2) Limited availability of testing kits and screening workstation creates roadblocks, especially in pandemic hotspots in economically challenged communities. Furthermore, early detection of COVID-19 is critical since COVID-19 induced pneumonia causes higher mortality rate in certain demographics. However, the effectiveness of early detection is further hindered mostly due to the inconsistent incubation period, which is the time between catching the virus and beginning to show symptoms that varies from 1-14 days. These challenges highlight the need to develop alternative approaches for COVID-19 detection.

Chest X-ray imaging is a frequently used modality for medical practitioners to assert or to deny the possibility of any pneumonia infection. A previous study identified that COVID-19 increases lung-density, which causes severe life threating Emphysema and chronic obstructive lung disease [7]. A COVID-19 chest X-ray image contains major image abnormalities such as horizontal white lines, bands, or reticular changes along with ground glass opacity [8]. Therefore, this imaging technique can be considered as a first-line screening tool to detect COVID-19 by exploring the persistent visual abnormalities in a chest X-ray image of COVID-19 infectant [9]. Although other imaging modalities, such as computed tomography (CT) provides higher resolution, chest X-ray image is cost-effective with high sensitivity [10,11]. Easy availability of X-ray machines also makes it an attractive choice for COVID-19 detection in the absence of testing kits and screening stations.

However, the biggest challenge of an X-ray based COVID-19 detection approach lies in manual examination of each X-ray image and extraction of the findings. This would require enormous time and presence of medical professionals. Thus, computer-aided chest X-ray examination methods are required for detection of COVID-19 cases from chest X-ray images. Towards this purpose, deep learning methods have proven to be useful in delivering high-quality results in addition to other advantages such as: (1) maximum utilization of unstructured data, (2) elimination of additional cost, (3) reduction of feature engineering, and (4) removal of explicit data labelling. Therefore, deep learning methods are often used to extract relevant features to classify image objects using its autonomous nature. Indeed, deep learning techniques have contributed significantly to analysis of medical images and achievement of excellent classification performance with less time-consuming simulated tasks [3].

In recent years, the use of deep learning methods in building convolutional neural networks (CNNs) has led to many breakthroughs in various computer vision-oriented research work such as image segmentation, image recognition and object detection. Previous research related to COVID-19 detection used various pre-trained CNN models such as VGG19, MobileNet, ResNet, and others for multi-class and binary classification task. For example, a combination of VGG19 and MobileNet in a multi-class classification study gave 97.8% accuracy in COVID-19 detection from healthy and pneumonia patients [12]. Another similar multi-class classification study using DarkCovidNet achieved a lower accuracy of 87% [13]. Using a binary classification system involving different ResNet models to classify COVID-19 versus non COVID-19 patients yielded higher accuracy of >98% [3,14].

Here, we evaluated the effectiveness of various pre-trained models and compared their performance with a CNN model developed here, DLH_COVID, in terms of detecting COVID-19 induced pneumonia cases from a set of chest X-ray images. To achieve this purpose, we first trained multiple pre-trained models and DLH_COVID using 80% train dataset. Then we validated each model using a 10% validation dataset. Finally, we selected the best model and performed accuracy check using a 10% test dataset. This analysis using a ‘training-validation-testing’ approach showed that DLH_COVID outperformed other models based on accuracy in COVID-19 detection from X-ray images. Furthermore, we repurposed the DLH_COVID model to develop a web-based user application that would benefit medical professionals to distinguish between COVID-19 and bacterial/viral pneumonia cases from chest X-ray images in a rapid and efficient manner.

## 2. RESULTS

### 2.1 Segregation of image dataset into train, validation, and test

We used a publicly available COVID-19 chest X-ray image repository [15] to obtain 6,432 images categorized into three groups: COVID-19, pneumonia and normal/healthy. These data included X-ray images with confirmed COVID-19, confirmed common pneumonia, and normal\healthy individuals. This dataset comprised 80% train dataset and 20% test dataset. To further prepare the test dataset for classification exercise, we applied stratified resampling method to split the test dataset into two subsets: 10% validation and 10% test subset. The 10% validation subset was used to prevent model overfitting and enhance model evaluation process. **Table 1** shows the final number of images distributed in the 80% train, 10% validation, and 10% test dataset used for the pre-trained model and DLH_COVID model development described below.

**Table 1.** Number of images segregated in the train, validation and test folders of the original dataset. Final distribution comprised 80% train, 10% validation and 10% test.

| Classification | Train | Validation | Test | Total |
| --- | --- | --- | --- | --- |
| COVID-19 | 460 | 58 | 58 | 576 |
| Normal | 1266 | 158 | 159 | 1583 |
| Pneumonia | 3418 | 427 | 428 | 4273 |
| **Total** | **5144** | **643** | **645** | **6432** |

In general, COVID-19 images look whiter compared to other images due to increased lung-density. Depending on the severity of pneumonia, some lung markings are partially obscured by the increased whiteness, which refers to as ground glass opacity. In more severe cases, the lung markings of COVID-19 images are completely removed due to whiteness, which is known as consolidation. Therefore, our artificial intelligence (AI) model considered radiographic appearance of multifocal ground glass opacity, horizontal white lines and consolidation as the determining factors for detecting COVID-19 from chest X-ray images of the infected patients [8]. **Figure 1** shows representative chest X-ray images of all the three conditions of COVID-19, pneumonia, and normal/healthy conditions.

**Figure 1.**
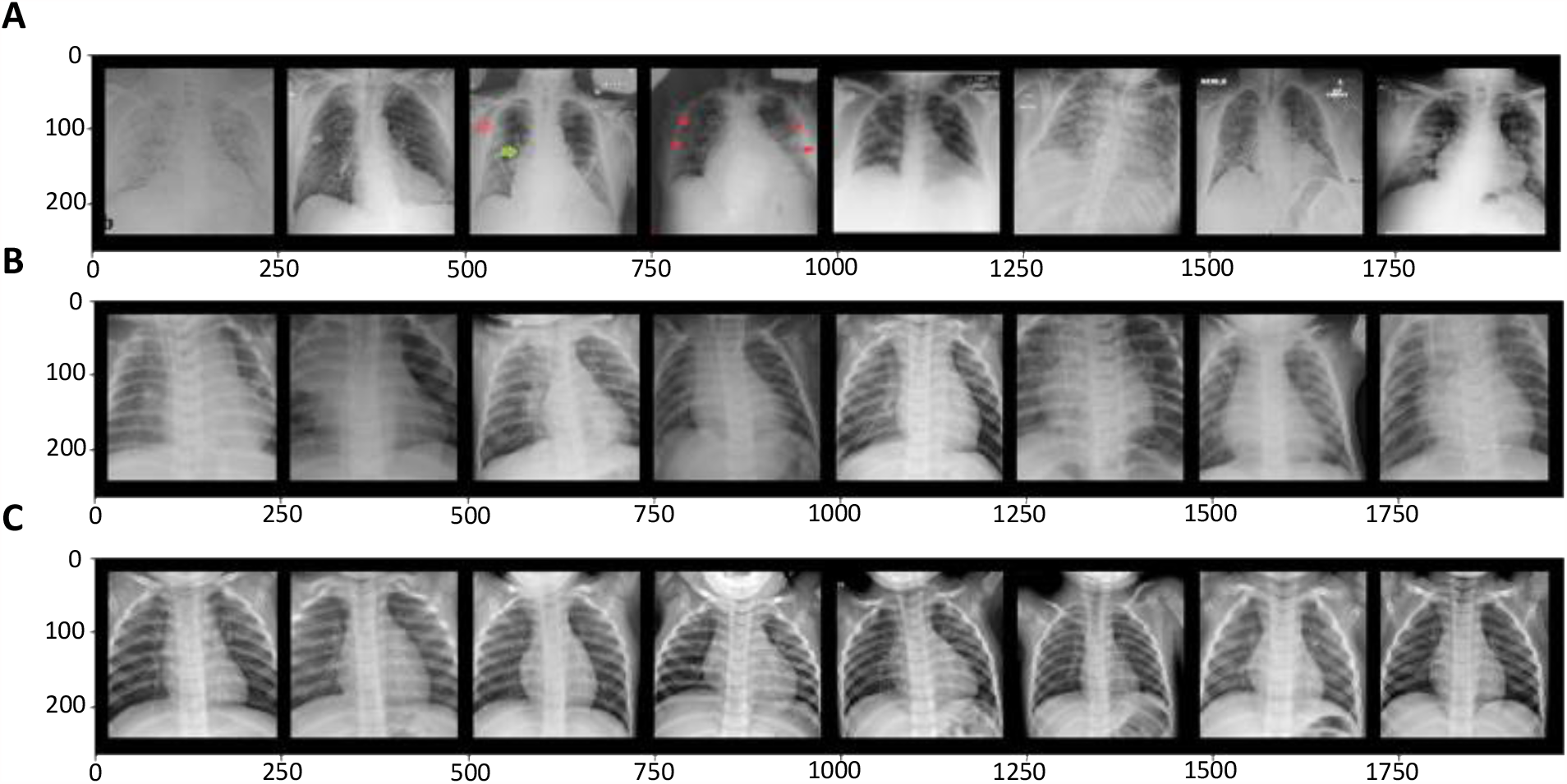
Representative chest X-ray images of (A) COVID-19, (B) pneumonia and (C) normal/healthy conditions. Note the increased ground glass opacity in COVID-19 X-ray images. Each image is of 224 × 224 resolution.

### 2.2 Pre-trained model selection

Previous research helped us to identify pre-trained models with high accuracy of COVID-19 detection from chest X-ray images [3,12,13,14]. These are the following models we used:

1. ResNet: These architectures were proposed by He et al. from Microsoft [16]. ResNet architectures introduced the use of residual layers and skip connection to solve the problem of vanishing gradient that may impact the weightage change in neural network.
2. VGG: These architectures were introduced by Oxford University’s Visual Geometry Group [17], where they demonstrated that using small filters of size 3 × 3 in each convolutional layer throughout the network may result in better performance. The main idea behind VGG architecture is that multiple small filters can make design simpler and reproduce similar results compared to that of larger filters.
3. GoogleNet: The main feature of GoogleNet/Inception architecture [18] is the innovation of inception module, which is a series of 1-by-1 convolutional layers/blocks used for dimensionality reduction and feature aggregation. This model comprised total 22 layers with 9 inception modules.
4. EfficientNet: This is a convolutional network with scaling method that uniformly scales width, depth, and resolution with a set of fixed coefficients. The base architecture is based on MobileNetV2, in addition to squeeze and extension blocks [19].

Furthermore, all the above pre-trained models are aligned with the transfer learning method, which we used to re-evaluate their performance in detection of COVID-19 from X-ray images.

### 2.3 DLH_COVID architecture

In this study, we developed a new convolutional neural network (CNN) for image classification exercise. We tested more than 100 different architectures with (i) different number of layers, (ii) different combination of number of neurons per layer, (iii) normalizations, (iv) max pooling techniques and (v) hyperparameters. We selected the one that achieved the best validation accuracy during training phase and named it as ‘DLH_COVID’. DLH stands for ‘Deep Learning in Healthcare’ and represents an artificial intelligence (AI) model purposed for COVID-19 detection. DLH_COVID consist of three convolutional layers followed by two fully connected linear layers. We used the 80% train dataset to carry out the aforementioned testing and finalized the DLH_COVID architecture. **Figure 2** shows the detailed architecture of DLH_COVID as described below:

**Figure 2.**
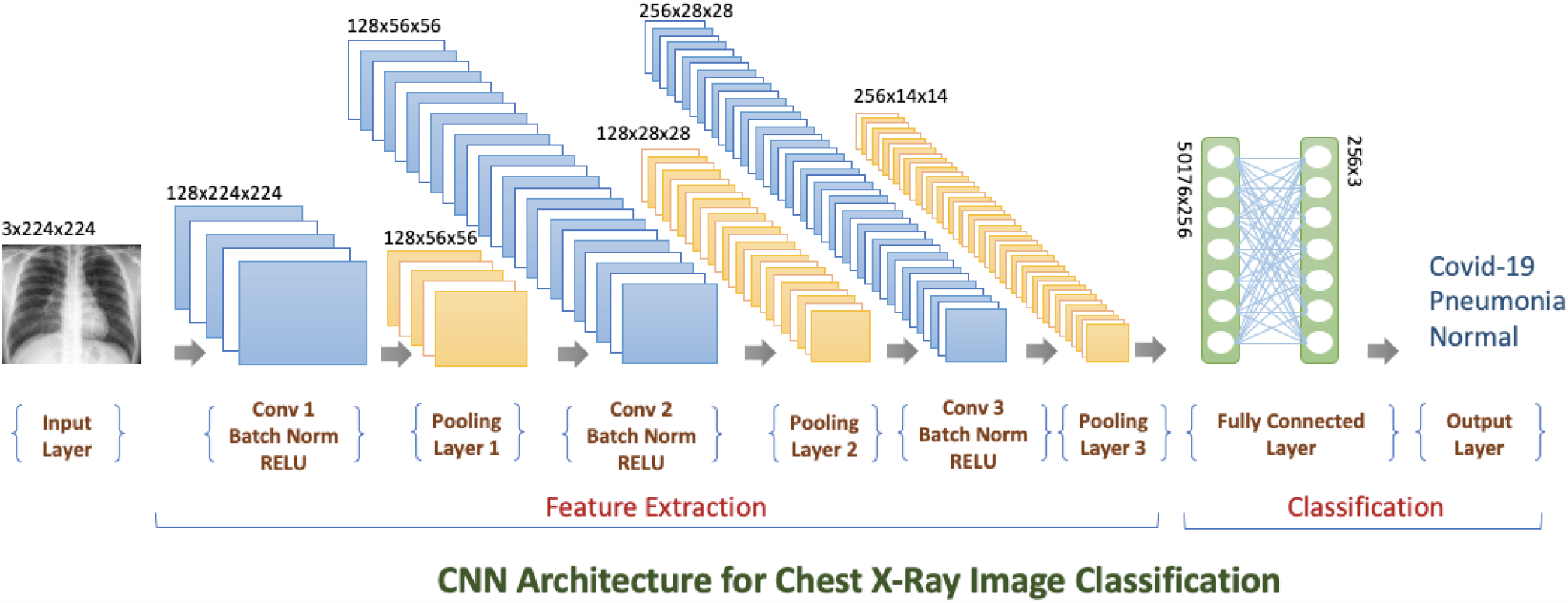
Schematic of DLH_COVID model architecture. It consists of three convolutional layers, two fully connected linear layers and additional intermediate maxpool layers. Input dimension of the CNN network: *3 × 224 × 224* and output dimension of the CNN network: *256 × 3*.

#### (1) First convolutional layer

the first convolutional layer has an input channel of dimension 3 to make it compatible with RGB image format. The kernel size was chosen to be of size 3 × 3 with stride of 1. We further applied image padding of size 1 to keep uniformity between input and output feature map. The output feature map at this layer is *128* × *224* × *224*.

Using batch normalization function (BatchNorm2d) followed by ReLU activation function in the intermediate layer, we normalized the feature map and incorporated non-linearity to the neural network. To reduce the number of training parameters and control the overfitting issue, a max pooling layer with kernel size 4 × 4 and stride 4 was introduced after ReLU activation function. The max pooling layer downsampled the feature map to *128* × *56* × *56*.

#### (2) Second convolutional layer

similar to the first layer, we added kernel size and image padding of size 3 × 3 along with stride of 1 to avoid any transformation in the dimension of the output feature map. The output feature map at this layer is *128* × *56* × *56*.

Like the previous intermediate layer between first and second convolutional layer, both batch normalization and ReLU activation function were applied sequentially to stabilize the neural network for further forward propagation. We introduced another max pooling layer with kernel size 2 × 2 and stride 2 to downsample the feature map to *128* × *28* × *28*.

#### (3) Third convolutional layer

the third convolutional layer was introduced to upgrade the output channel. Further, we incorporated another set of maxpool and ReLU activation layers, which transformed the output feature map to *256* × *14* × *14*. This map was fed into the linear layer in the later stage of feature extraction.

#### (4) Fully Connected Layer

Finally, two connected linear layers in conjunction with a dropout layer were added to reduce data overfitting and streamline the final output channel. We used recommended dropout value of p = 0.5 [20].

A flattened version of the feature map was passed to the first fully connected linear layer. So, the input dimension of the first linear layer consisted of *256* × *14* × *14 = 50176 nodes* and later it was integrated with the final linear layer of *256 nodes*. The final output dimension of the DLH_COVID architecture was 3 corresponding to the total number of image classes: COVID-19, pneumonia and normal/healthy.

### 2.4 Image pre-processing

Previous research has shown that downsampling input images to a lower resolution increased effectiveness of CNN classification models [3]. Therefore, we rescaled all train, validation, and test images into standard size 224 × 224 to maintain uniformity in image resolution. Since one of the selected pre-trained models, Inception-V3, is only compatible with resolution 299 × 299 [21], we made further readjustments to image resolution. The pre-processing step also included center crop mechanism, which was applied on all the X-ray images to reduce background noise and enhance focal length position. In addition, following image augmentation techniques on 80% train dataset during data pre-processing step [3] was applied:

(i) Rotation: rotated images at various angles between -10^0^ to +10^0^ and added these augmented images in the train dataset, (ii) Gaussian Blur: applied Gaussian filter of kernel size (3,3) on the images to remove high frequency components, (iii) Flip: images were randomly flipped horizontally and vertically to achieve data augmentation, (iv) RandomResizecrop: images were cropped and resized randomly to overcome any unforeseen data overfitting issues, and (v) Grayscale transformation: images were transformed from RGB (three channels) to grayscale (one channel). However, none of the image augmentation techniques were effective during training phase as the augmented images incurred high train and validation loss. Therefore, we used only non-augmented images that was necessary to eradicate potential pitfall of the classification exercise in later stages.

### 2.5 Model optimization

In the model training phase, we relied on both manual intervention and dynamic approach to fine-tune all model hyperparameters such as (i) learning rate, (ii) number of epochs and (iii) optimizer. We focused on determining the optimal learning rate, which is an essential hyperparameter as it correlates with the loss function. However, the selection of optimal learning rate can be cumbersome because large learning rate can cause weights of the neurons to converge quickly, or smaller learning rate may cause delays in training the network [3]. Therefore, we followed two approaches: (i) train model with static learning rate, often in the range between small and default learning rate le-2 and (ii) adopt dynamic learning rate using scheduler functionality of PyTorch to ascertain the final optimal learning rate.

Initially each model was separately trained using two fixed learning rate le-3 and le-2, and the model with learning rate le-3 inclined towards achieving better performance. We observed that the pre-trained model exhibited more stable validation accuracy with lower number of epochs ranging between 20 and 25. On the contrary, the DLH_COVID model required more epochs with fixed learning rate to achieve stable validation accuracy. **Figure 3A** shows the validation and train losses of our CNN model using a fixed learning rate of le-3 and 100 epochs. The line plot also shows a steep increase in validation loss at epoch 25 and gradually increasing gap between train and validation loss based on higher number of epochs. To avoid inconsistencies between train and validation loss, we adopted AutoML dynamic learning rate technique using scheduler functionality of PyTorch to train the DLH_COVID model. In this scenario, the model training started with a learning rate of 1e-3 and gradually reduced to 1e-5. Although we defined maximum 100 epochs for training, we noticed that the validation loss of the DLH_COVID model did not improve after 30 epochs. For instance, the validation accuracy varied from 0.92 to 0.97 when we trained the model with a fixed learning rate and 25 epochs. However, it remained stable between 0.96 and 0.97 when we adopted variable learning rates and epochs. **Figure 3B** shows that the train and validation loss appeared to be more stable after incorporating dynamic learning rate procedure.

**Figure 3.**
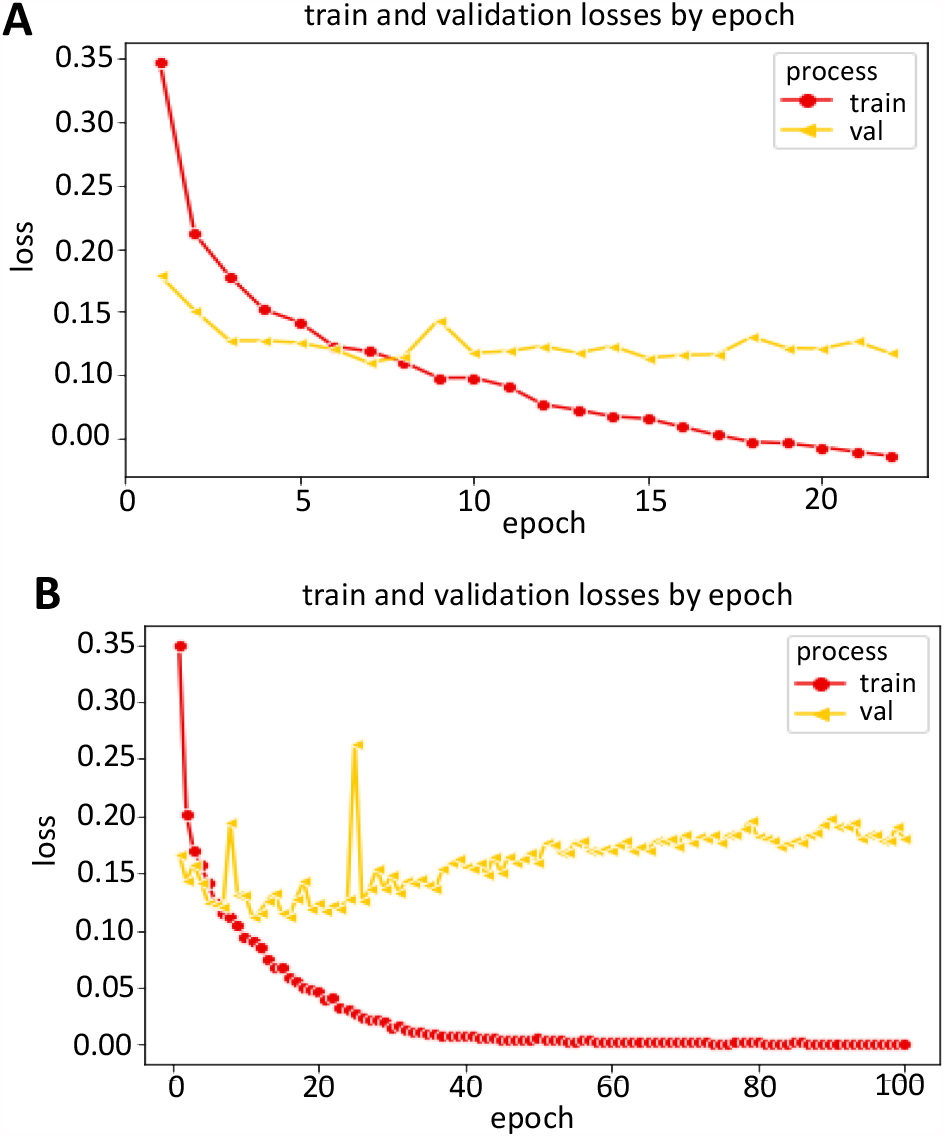
Train and validation loss with (A) fixed learning rate over 100 epochs, and (B) dynamic learning rate over 25 epochs of DLH_COVID model.

Taken together, the dynamic learning rate technique was proven to be beneficial to determine the optimized learning rate and number of epochs for the DLH_COVID model, but a similar dynamic approach was not necessary for the pre-trained models. Furthermore, we used both Adam and SGD optimizer to train each model sequentially. Although training CNN model with Adam optimizer is less time consuming in comparison to training with Stochastic Gradient Descent (SGD) optimizer, all the models yielded higher accuracy by using the latter optimizer. We assumed that this scenario occurred due to the failure of Adam optimizer to converge to an optimal point under specific settings [22]. Therefore, SGD was selected as the primary optimizer for this study.

### 2.6 Performance evaluation

Model performance was evaluated using the common statistical measure confusion matrix from which we obtained various metrics like accuracy, precision, recall and f1-score. In Table 2, we show the evaluation metrics of both pre-trained models and DLH_COVID based on 10% validation dataset. We considered accuracy score as the best statistical measure to compare performance of the pre-trained models with that of the DLH_COVID model. Initial analysis of validation data revealed the following: (1) none of the pre-trained models achieved more than 96% accuracy score (**Table 2**); (2) DLH_COVID model with a simpler architectural design was able to outperform these pre-trained models in terms of detection of COVID-19 from 10% image validation dataset.

**Table 2.**
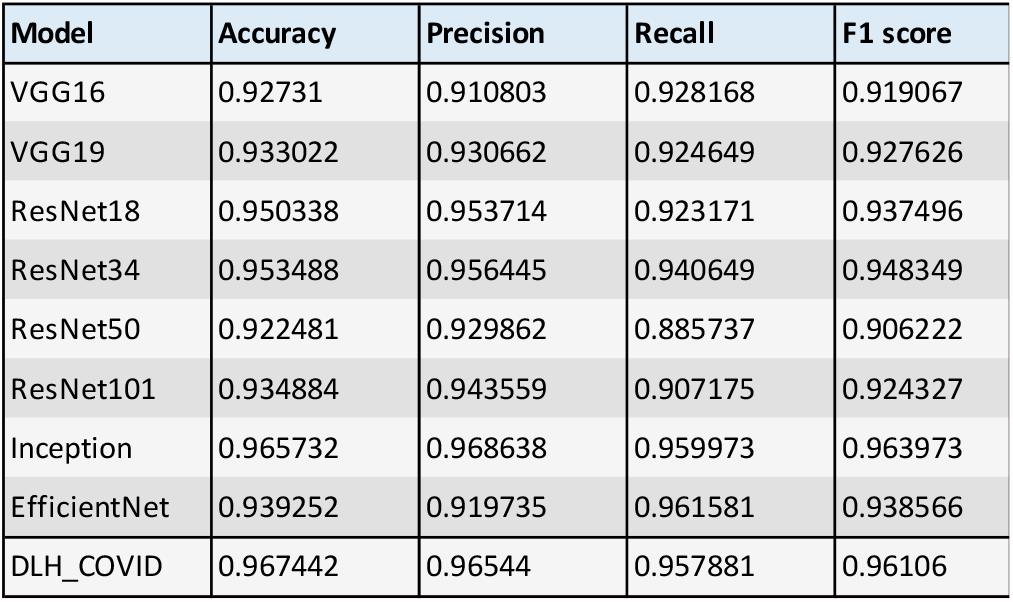
depicts the preliminary evaluation metrics of different models acquired for this image classification task. We used ‘accuracy’ column values to measure performance of each model and selected DLH_COVID, since it exhibited the highest accuracy.

To support the results obtained from performance evaluation shown in Table 2, we carried out k-fold cross validation procedures. This was done to evaluate generalized performance of the DLH_COVID model and perform accuracy score comparison with other pre-trained CNN models. For this purpose, we selected commonly used variation of folds, that is, k = 5. **Figure 4** depicts the five-fold cross validation where one unique fold represents test dataset, and the remaining folds are chosen as training dataset. The models were fitted on the training dataset and validated on the test dataset. The key performance measures of accuracy score, precision, recall and f1-score obtained for each of the fold were averaged to produce the final five-fold performance metrics of the model.

**Figure 4.**
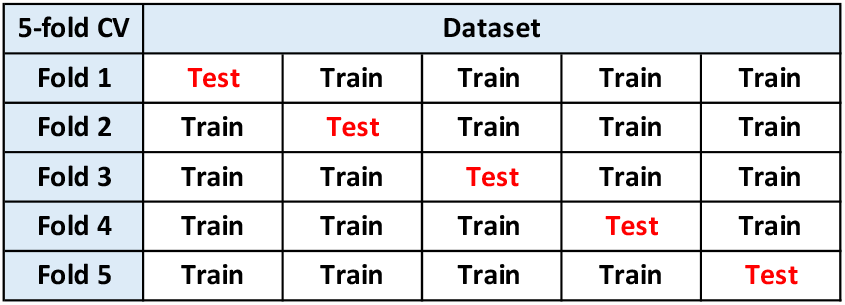
Schematic depiction of 5-fold cross validation technique. One unique fold represents test dataset, and the remaining folds are chosen as training dataset.

During the training phase, the weights of our neural network model were reset at the beginning of each fold run to avoid data overfitting issue. However, that technique was not necessary for other models because only last layer of the pre-trained network was fitted during execution of each fold. The training parameters for all the pre-trained networks were as follows: learning rate: le-3, batch size = 32, number of epochs = 20. We picked the best VGG and ResNet model based on previous validation results (Table 2) for the cross-validation exercise. **Table 3** shows that DLH_COVID exhibited best performance by achieving highest average accuracy score of 95% on the one-fold test dataset.

**Table 3.** shows the 5-fold cross validation results of pre-trained and DLH_COVID model. For this model evaluation process, we selected the pre-trained models that exhibited higher accuracy value in the previous validation stage. DLH_COVID model yielded highest accuracy here as well.

| MODELS | Performance results (%) |  |  |  |  |
| --- | --- | --- | --- | --- | --- |
|  | Fold | Accuracy | Precision | Recall | F1-score |
| VGG19 | Fold-1 | 92% | 91% | 89% | 90% |
|  | Fold-2 | 92% | 91% | 92% | 91% |
|  | Fold-3 | 93% | 94% | 92% | 93% |
|  | Fold-4 | 94% | 94% | 92% | 93% |
|  | Fold-5 | 91% | 90% | 89% | 90% |
|  | Mean | 92% | 92% | 91% | 91% |
| RESNET34 | Fold-1 | 94% | 93% | 93% | 93% |
|  | Fold-2 | 93% | 92% | 94% | 93% |
|  | Fold-3 | 96% | 97% | 95% | 96% |
|  | Fold-4 | 94% | 96% | 93% | 94% |
|  | Fold-5 | 95% | 95% | 95% | 95% |
|  | Mean | 94% | 94% | 94% | 94% |
| DLH_COVID | Fold-1 | 95% | 93% | 92% | 92% |
|  | Fold-2 | 96% | 95% | 94% | 94% |
|  | Fold-3 | 95% | 95% | 93% | 94% |
|  | Fold-4 | 96% | 97% | 93% | 95% |
|  | Fold-5 | 94% | 96% | 89% | 92% |
|  | Mean | 95% | 95% | 92% | 93% |
| INCEPTION | Fold-1 | 90% | 90% | 84% | 87% |
|  | Fold-2 | 89% | 88% | 90% | 89% |
|  | Fold-3 | 90% | 91% | 87% | 88% |
|  | Fold-4 | 92% | 94% | 88% | 91% |
|  | Fold-5 | 90% | 88% | 88% | 88% |
|  | Mean | 90% | 90% | 87% | 88% |

### 2.7 Final model accuracy check

DLH_COVID model attained highest accuracy score in comparison to other pre-trained models in detection of COVID-19 images from 10% validation dataset. Moreover, it achieved almost similar performance in baseline k-fold cross validation process. This further demonstrated the robustness of the model in classifying X-ray images. Therefore, we decided to utilize this AI model for performing final validation on 10% test dataset and report the accuracy in a form of confusion matrix. **Figure 5** shows the 3 × 3 confusion matrix where each row and column represent one image class. The black cells of the matrix signify the number of images that were evaluated as False Positive and False Negative images. Taken together, a total of 21 out of 645 test images were misclassified by the DLH_COVID model, and the majority of misclassification occurred between the X-rays of pneumonia and normal image class.

**Figure 5.**
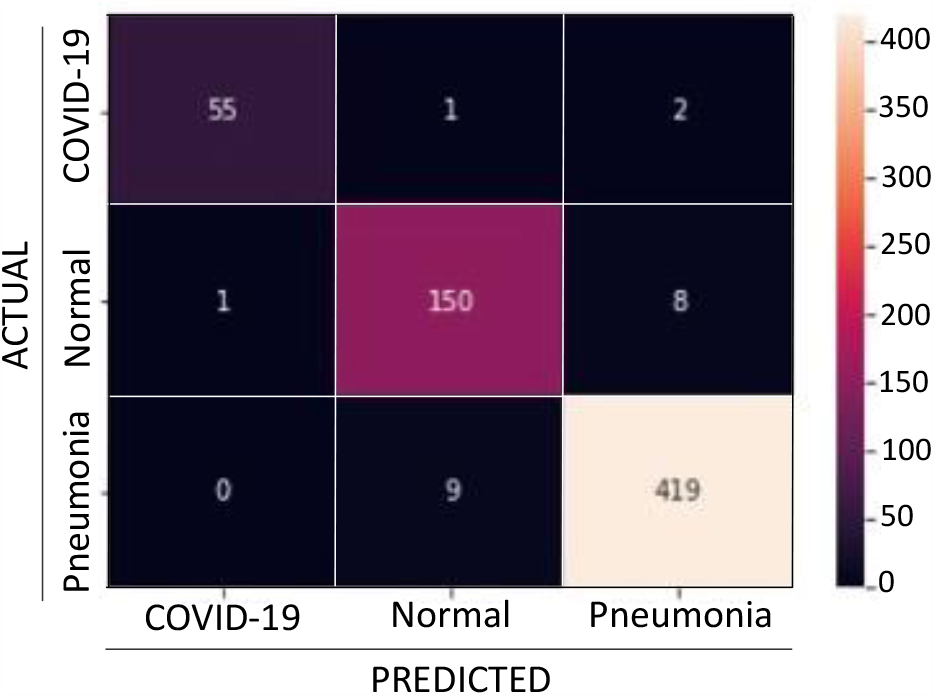
Confusion matrix of DLH_COVID model. Each entry of the matrix denotes the number of predictions made by the model where it was classified correctly or incorrectly. All the entries located in black cells represent False Positive or False Negative value of the matrix.

### 2.8 COVID-19 detection web-based application

Lastly, we integrated the DLH_COVID model with a cloud-based web platform, which is easily accessible and easily shareable with medical professionals around the world including pandemic hotspots. This AI system is a browser-based web application (link: https://toad.li/xray) that takes chest X-ray image as input data, processes image data through the DLH_COVID classification model and generates probability score of the image class as output. The image class corresponds to COVID-19, pneumonia and normal/healthy condition. Since the probability score is a measure to determine the class of the uploaded chest X-ray image, the image class with the highest score is predicted as the final output of the AI model. For example, **Figure 6** shows that DLH_COVID was able to accurately predict the presence of COVID-19 from an uploaded image. It estimated a high probability score >99% in the image class of COVID-19. Similarly, AI model enables users to detect bacterial/viral pneumonia from chest X-ray images as well, demonstrating that the model can execute multi-class image classification in real time.

**Figure 6.**
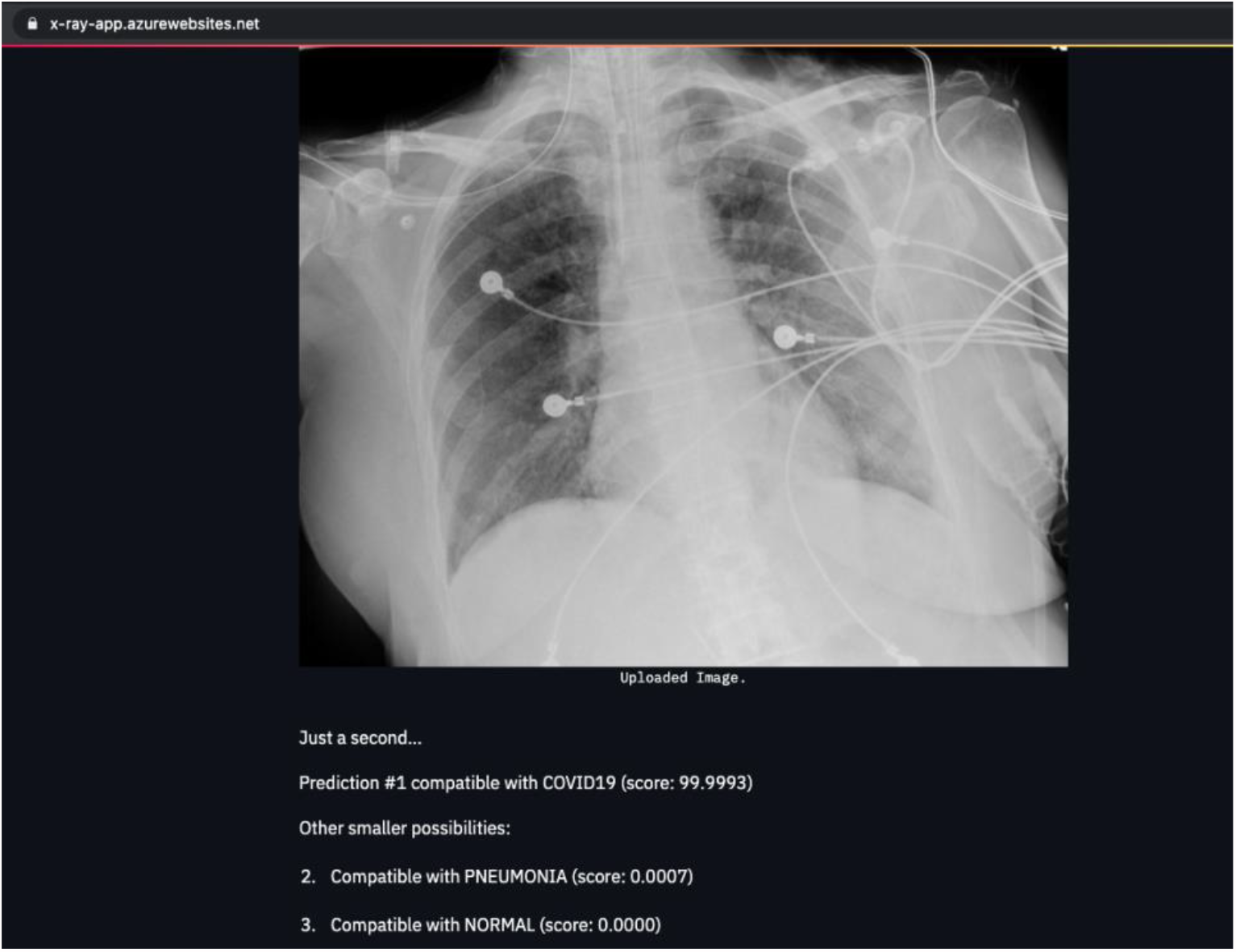
Sample screenshot image of COVID-19 detection web application. A chest X-ray image of a COVID-19 sample was uploaded as input. Within a few seconds, the application provided an output prediction of COVID-19 with 99.9993 probability score. In contrast, probability scores of 0.0007 and 0.0000 was obtained for pneumonia and healthy X-ray samples, respectively.

## 3. DISCUSSION

The accurate detection of COVID-19 cases in a timely manner has become a salient factor to curb the outbreak in pandemic hot spots. Recent research studies have shown the usefulness of chest X-ray images in COVID-19 diagnosis [23,24,25,26,27]. Our research work augments these previous studies by reinforcing the importance of chest X-ray imaging in similar medical diagnosis procedures. We also showcase an AI system driven by a DLH_COVID model established in this study, which can predict the presence of COVID-19 in a chest X-ray image and distinguish it from pneumonia or normal condition. It also broadens the scope of classification levels by additionally detecting pneumonia cases with a reasonable high accuracy score from X-ray images.

The DLH_COVID model is unique, reliable and independently developed without any influence of the transfer learning method. We attempted to fine-tune the model through trial-and-error approach, as theoretically it is impossible to determine the optimal hyperparameters without going through a comprehensive series of training cycles. The experimental results during prospective validation phase suggests that the DLH_MODEL outmatched most of the pre-trained models since it showed a promising accuracy of 96% in classifying COVID-19, pneumonia, and normal/healthy cases from the image dataset. Furthermore, k-fold cross validation results reassured the efficacy of the DLH_COVID model in determining multiple image classes without performing any explicit data cleaning process. Given the simple CNN architecture of the DLH_COVID model, it can be easily extended by incorporating additional convolutional layers or by readjusting other model components. Therefore, we believe that AI researchers could easily leverage this model through transfer learning or ensemble learning method for future studies related to similar medical image classification.

However, there were 21 images misclassified by the DLH_COVID model during the final accuracy test. This is primarily due to lower image resolution, which prevented the AI model to predict image class accurately. Furthermore, we encountered data scarcity issue throughout the study as the model was not trained with a sufficient number of COVID-19 images in comparison to that of other image classes. This limited availability of training data pertaining to a given image label created class imbalance constraint [28], which often causes model overfitting issue in neural network. Although we added dropout layer to DLH_COVID architecture and executed k-fold cross validation to overcome this issue, this is not sufficient to alleviate the overfitting issue and make the AI model more generalized to handle unseen test data. We believe that model sustainability and accuracy can be drastically improved by incorporating a larger COVID-19 chest X-ray image dataset. In addition, the AI model can be further improved by enhancing the quality of the misclassified images using computer vision techniques such as stretching, slicing and histogram equalization [29].

The recent success of an AI system in carrying out similar X-ray image classification task supplements our objective of developing a user interface system driven by DLH_COVID model [30]. On the contrary, another recent study revealed that applicability of deep learning models in real hospital management ecosystem is still unclear [6]. Thus, it is imperative for more assessments to be made to assert the reliability of AI systems as an important tool for COVID-19 diagnosis. Therefore, we have ensured that the DLH_COVID based web application is publicly accessible so that doctors and radiologists can easily test the underlying AI model in clinical settings and capture results accordingly. Critical feedback from medical professional will provide additional guidance to improve the DLH_COVID AI model. This will eventually benefit COVID-19 clinical management settings in pandemic hotspots with an accurate and fast diagnosis process in the foreseeable future.

## 4. METHODS

### 4.1 Pre-trained model reusability

Transfer learning method was employed to demonstrate the reusability of pre-trained models that had shown good performance in carrying out COVID-19 X-ray image classification task in previous research [3]. Transfer learning was used for the following reasons: (a) since the initial layers of the network are already trained, it saves train time and avoid huge computational power required for training, (b) requires low volume of training data given the lack of large numbers of COVID-19 X-ray images, and (c) proven to be beneficial to fit training dataset comprised of standard size images. We used VGG16, VGG19 [17], ResNet18, ResNet34, ResNet50, RestNet101[16], Inception [18] and EfficientNet [19] that are aligned with the transfer learning method.

### 4.2 DLH_COVID model development

We used a conventional CNN architecture to develop a new image classification model DLH_COVID.

### 4.3 Dataset used

We used a publicly available COVID-19 chest X-ray images repository [15] to obtain total 6,432 images categorized into three groups: COVID-19, pneumonia and normal/healthy. These data included images with confirmed COVID-19, confirmed common pneumonia, and normal\healthy individuals. This dataset comprised 80% train dataset and 20% test dataset.

### 4.4 Model Optimization

We use hyperparameter optimization as it helped us to determine the optimal combination of hyperparameters that minimizes the train and validation loss during the model training phase. The following methods given below were proven instrumental to achieve the purpose of model optimization.

#### (1) Learning rate scheduling method

We used PyTorch library module optim() that provides several methods to adjust learning rate based on the number of epochs during model training phase [31]. This functionality benefits the model by reducing the learning rate dynamically if no improvement is seen for a fixed number of epochs.

#### (2) SGD optimizer

Stochastic Gradient Decent (SGD) is a variant of gradient decent algorithm which allows the optimizer to compute on a small subset or random selection instead of the whole training dataset. SGD performs computation much faster as it removes redundancy by performing one model hyperparameter update at a time [32].

#### (3) Adam optimizer

Adaptive Moment Estimation (Adam) is an adapting learning rate algorithm mainly purposed for gradient-based optimization of stochastic objective functions. This optimizer method is very straightforward to implement, computationally efficient, less time-consuming and requires little memory to process [32].

#### 4.5 Performance metrics

Model performance was evaluated using the common statistical measure confusion matrix from which we obtained various metrics like accuracy, precision, recall and f1-score. These measures are briefly described below.

(i) Accuracy: a parameter that evaluates the correctness of the model by measuring a ratio of accurately predicted cases out of total number of cases. Mathematical formula is expressed as:

*Accuracy = (TN + TP) / (TN + TP + FN + FP)* [33, 34]

True Positive (TP): number of correctly identified COVID-19/pneumonia X-ray images,

False Negative (FN): number of incorrectly classified COVID-19/pneumonia X-ray images,

True Negative (TN): number of correctly identified healthy X-ray cases.

False Positive (FP): number of incorrectly identified healthy X-ray cases.

(ii) Precision: the ratio of correctly predicted positive cases to the total predicted positive cases. High precision relates to a low false positive rate. It is expressed as:

*Precision = TP / (TP + FP*) [35]

(iii) Recall: It is the ratio of correctly predicted positive observations to all observations in actual class.

*Recall = TP / (TP + FN)* [36]

(iv) F1-Score: F1 Score is measured in case of uneven class distribution especially with a large number of true negative observations. It provides a balance between Precision and Recall.

*F1-score = 2 × [(Precision x Recall) / (Precision + Recall)]* [37]

In addition, we performed a k-fold cross validation to support the initial performance metric evaluation.

### 4.6 Software and infrastructure

All the models were trained using open-source machine learning library PyTorch [38]. To harness better computing power and maximum throughput, we used both Google Colab cloud platform and machines with modern GPU configurations to train the CNN models. We used Microsoft Azure’s cloud service to deploy DLH_COVID model and made it accessible via web browser.

## Data Availability

The chest X-ray image dataset used for this research is available at publicly accessible Kaggle website.

https://www.kaggle.com/prashant268/chest-xray-covid19-pneumonia

## DATA AVAILABILITY

The chest X-ray image dataset used for this research is available at https://www.kaggle.com/prashant268/chest-xray-covid19-pneumonia.

## CODE AVAILABILITY

The python code of the DLH_COVID model network along with the model hyperparameters required to reproduce the image class predictions is available at https://github.com/soudey123/COVID-19-CHEST-X-RAY-IMAGE-CLASSIFICATION_UIUC

## ACKNOWLEDGEMENTS

We thank Prashanth Patel for creating the COVID-19 dataset and making it publicly accessible on the Kaggle website. We thank Debosmita Sardar, PhD for providing feedback during manuscript preparation. We thank Dr. Jesamar Mattos for providing feedback on the efficacy of the DLH_COVID model during final accuracy check. We thank our Professor Jimeng Sun, PhD for giving us the opportunity to work on this project.

## AUTHOR CONTRIBUTIONS

Gunther Correia Bacellar: Conceptualization; data research; DLH_COVID model architecture design, hold-out validation, and deployment; data visualization; review and edit manuscript.

Mallikarjuna Chandrappa: Pre-trained model research, development and hold-out validation, data visualization; review and edit manuscript.

Rajlakshman Kulkarni: Pre-trained model research, development and hold-out validation, data visualization; review and edit manuscript.

Soumava Dey: Conceptualization; literature survey; DLH_COVID model architecture design, hold-out and k-fold cross validation; write and edit manuscript; overall project management.

